# Loneliness, physical activity and mental health during Covid-19: a longitudinal analysis of depression and anxiety between 2015 and 2020

**DOI:** 10.1101/2020.07.30.20165415

**Authors:** Byron Creese, Zunera Khan, William Henley, Siobhan O’Dwyer, Anne Corbett, Miguel Vasconcelos Da Silva, Kathryn Mills, Natalie Wright, Ingelin Testad, Dag Aarsland, Clive Ballard

**Affiliations:** University of Exeter Medical School, College of Medicine and Health, UK; Department of Old Age Psychiatry, Institute of Psychiatry, Psychology and Neuroscience, King’s College London, UK; Global Public Health, Public Health England, London, SE1 8UG, UK; Centre for Age-related Medicine - SESAM, Stavanger University Hospital, Stavanger, Norway

## Abstract

**Background:** Loneliness and physical activity are important targets for research into the impact of COVID-19 because they have established links with mental health, could be exacerbated by social distancing policies and are potentially modifiable.

**Method:** We analysed mental health data collected during COVID-19 from adults aged 50 and over alongside comparable annual data collected between 2015 and 2019 from the same sample. Trajectories of depression (PHQ-9) and anxiety (GAD-7) were analysed with respect to loneliness, physical activity levels and a number of socioeconomic and demographic characteristics using zero-inflated negative binomial regression.

**Results:** 3,281 people completed the COVID-19 mental health questionnaire, all had at least one data point prior to 2020. In 2020, the adjusted PHQ-9 score for loneliness was 3.2. (95% CI: 3.0-3.4), an increase of one point on previous years and 2 points higher than people not rated lonely, whose score did not change in 2020 (1.2, 95% CI: 1.1-1.3). PHQ-9 was 2.6 (95% CI: 2.4-2.8) in people with decreased physical activity, an increase of 0.5 on previous years. In contrast, PHQ-9 in 2020 for people whose physical activity had not decreased was 1.7 (95% CI: 1.6-1.8), similar to previous years. A similar relationship was observed for GAD-7 though the differences were smaller and the absolute burden of symptoms lower.

**Conclusions:** After accounting for pre-COVID-19 trends, we show that experiencing loneliness and decreased physical activity are risk factors for worsening mental health during the pandemic. Our findings highlight the need to examine policies which target these potentially modifiable risk factors.

## Introduction

In order to contain and reduce the spread of COVID-19, the UK government introduced lockdown measures on 23^rd^ March 2020 which restricted time permitted outside and all non-essential in-person contact. Those with certain high risk medical conditions were advised to ‘shield’ (i.e. not leave the house for 12 weeks) and those aged 70 and over were advised to strictly adhere to the restrictions. The potential mental health impacts of this have been highlighted in a number of high profile commentaries, with possible mechanisms including the pressures of lockdown, anxieties about infection and the knock-on economic consequences (1–7). Previous research into mental health in the pandemic has largely focused on socioeconomic, demographic and clinical comorbidities, with younger age, female gender and low socioeconomic status being consistently associated with higher risk (8–11). While these links are undoubtedly important, research must also focus on potentially modifiable risk factors.

Loneliness and physical activity are critical mediators of mental health and therefore warrant close consideration during the pandemic (5,12,13). The pandemic may lead to low activity levels and exacerbate the relationship between loneliness and mental health in some (for example through social distancing and movement restrictions) and as such they may represent modifiable targets for resilience and management programmes. Longitudinal data covering the pre-pandemic and pandemic period is needed to address this key question. Three representatively sampled surveys (two US and one UK) of data pre and during the pandemic reported no significant changes in loneliness but no links were drawn between the interactions with the pandemic on mental health levels (11,14,15). One cross sectional study linked loneliness with worse mental health and a second study indicated that people with low social support (a possible proxy for loneliness) had a more severe trajectory of depression during the pandemic (8,16). However without data prior to 2020, it is impossible to evaluate fully the specific importance of these factors during the pandemic, in particular whether these relationships reflect well-established links between loneliness and mental health or whether there was a specific effect of the pandemic. Though highlighted as important in commentaries, there has been little research into the links between physical activity and mental health during the pandemic; to our knowledge, the only published study used a cross sectional design (17).

To address the gap in research around the impact of loneliness and physical activity on mental health during COVID-19 we analysed data from 3,281 participants, all of whom had mental health data available from before the pandemic. We hypothesized that trajectories of depressive and anxiety symptoms in people who were lonely and whose physical activity had decreased during the pandemic would be adversely affected. In addition, we also examined a number of other demographic and socioeconomic variables on mental health trajectories.

## Method

### Study design and setting

The study was conducted with participants from the PROTECT study. PROTECT is a longitudinal study of mental and cognitive health with annual assessment in people over the age of 50 which was launched in November 2015 (http://www.protectstudy.org.uk/) (18). There are currently over 25,000 people enrolled in PROTECT. Written informed consent was obtained online from all participants.

In May 2020, around 4.5 years after PROTECT started, a specific COVID-19 mental health questionnaire was launched in PROTECT, again completed online. Here we present an analysis of data collected between 13^th^ May and 8^th^ June 2020, combined with existing data from previous years.

The authors assert that all procedures contributing to this work comply with the ethical standards of the relevant national and institutional committees on human experimentation and with the Helsinki Declaration of 1975, as revised in 2008. All procedures involving human subjects/patients were approved by the UK London Bridge National Research Ethics Committee (Ref: 13/LO/1578) and the COVID-19 mental health questionnaire was approved by the same committee (as an amendment) on 6^th^ April 2020.

### Participants

The PROTECT cohort includes people aged 50 or over at enrolment living in the UK. Additional inclusion criteria are: access to a computer and internet, able to read and write English and no diagnosis of dementia. All participants who opted in to study communications were invited to complete the COVID-19 mental health questionnaire.

### Variables

The principal outcome measure for this study was PHQ-9 and GAD-7 scores (measuring depression and anxiety respectively) record pre-pandemic (2015-2019) and during the pandemic (2020).

### PROTECT pre-pandemic data collection 2015-2019

Before the pandemic, all participants completed a series of online self-report questionnaires, which included demographic information (date of birth, gender, highest level of education [left school at 16, left school at 18, undergraduate degree, post-graduate degree], employment status [full time, part-time, self-employed, retired, unemployed] and marital status [married/civil partnership/co-habiting, widowed/divorced/separated, single], history of psychiatric illness). In addition, mental health assessments were completed annually prior to the pandemic as well as in 2020.

Depression was assessed with the PHQ-9, a 9 item questionnaire which assesses the frequency of depressive symptoms over a two week window (19). Each item is rated on a 4 point scale (0=not at all; 1=several days; 2=more than half the days; 3= nearly every day). Anxiety was assessed with the GAD-7, a 7 item questionnaire assessing the frequency of anxiety symptoms over a two week window (20). The ratings are the same as PHQ-9. For both scales, a threshold of 5 or above is indicative of mild and 10 or above is indicative of moderate or severe symptoms.

Participants completed up to four annual GAD-7 and PHQ-9 assessments spread over 5 years between 2015 and 2019 (depending on enrolment date). Enrolment to PROTECT is open continuously and started with a national publicity drive in October and November 2015, as a result the majority of current participants enrolled in those two months. For those who completed the COVID-19 mental health questionnaire this figure was 1,930 (59%). After the initial wave of enrolment, 405, 382, 338 and 18 enrolled in 2016, and in 2017, 2018 and 2019 respectively. Thus, most completed annual assessments between October and January of each year. PROTECT pre-pandemic data was available from a data-freeze in early October 2019.

### Mental health during COVID-19 May 13^th^-June 8^th^ 2020

Symptoms of COVID-19 infection. Participants were asked whether they had any of the main symptoms of COVID-19 in the last two weeks (which at the time were a new persistent cough for more than 24 hours or a high temperature) or if they had been hospitalized with COVID-19 in the last four weeks.

Physical activity changes. Participants were asked about changes in their physical activity since March 2020. The data were categorised to identify people who reported a decrease in their level of physical activity and those who did not.

Physical illnesses. Participants were asked if they had any of the following conditions associated with moderately increased risk of severe illness from coronavirus: long-term respiratory illness, chronic heart disease, chronic kidney disease, liver disease, neurological disease, diabetes, illness affecting the spleen, weakened immune system or BMI>=40. They were also asked if they had any of the following conditions which would require them to shield (high risk of severe illness from coronavirus): received an organ transplant and remain on ongoing immunosuppression medication, undergoing active chemotherapy or radiotherapy, cancer of the blood or bone marrow who are at any stage of treatment, severe chest conditions such as cystic fibrosis or severe asthma (requiring hospital admissions or course of steroid tablets), severe diseases of body systems. People were also asked if they had received a letter advising them to shield and if they answered yes they were included in the high risk group. These physical conditions were coded 0 (no relevant conditions); 1 (moderate risk conditions) and 2 (high risk conditions).

Loneliness. Loneliness was assessed using the three item UCLA loneliness scale (21). The questions ask how often the participant has felt lack of companionship, left out and isolated from others with the possible answers being ‘hardly ever’, ‘some of the time’ and ‘often.’ Loneliness was treated as binary for this analysis, dichotomized into those experiencing any loneliness (i.e. rating at least ‘some of the time’ on any question) and those experiencing none.

Finances. Participants were asked to respond yes or no the question “Has the COVID-19 (coronavirus) had had a negative impact on your finances?”

### Statistical methods

The statistical analyses were carried out in two stages.

In the full cohort we first undertook a case level analysis of PHQ-9 and GAD-7 rated in 2020 during the pandemic, categorizing both into a three level factor (see above for cut offs) representing no, mild and moderate-to-severe symptoms. Differences in the proportions of current depression and anxiety levels by risk factor were analysed using the chi-square test. We then undertook descriptive analysis of the change in case level proportions between 2019 and 2020.

For the second and principal analysis, we examined trajectories of PHQ-9 and GAD-7 between October 2015 (the start of the PROTECT study) and 8^th^ June 2020. Initial analysis of PHQ-9 and GAD-7 total scores using linear mixed effects models showed evidence of departure from the assumption of normally distributed residuals (see supplement). This could not be rectified by transformations and instead we considered models for counts of symptoms. A zero-inflated negative binomial regression (ZINB) was chosen for each scale due to over-dispersion and evidence of excess zeros. Zero inflated models use a mixture model approach in which the population is assumed to consist of an at-risk subgroup, and a sub-group not at risk for PHQ-9 and GAD-7 symptoms during the study period (the source of the excess zeros). The model is comprised of two components: the first accounts for the distribution of symptoms in the at-risk population (negative binomial component), the second is a logit model accounting for factors associated with membership of the non-risk sub-population (zero-inflated component). A random intercept term was included to allow for correlations between repeated measurements on the same individual. The Vuong test statistic was used to compare the fit of zero-inflated models with single-equation Poisson and negative binomial models.

Univariate models were run for both PHQ-9 and GAD-7 for loneliness and physical activity, as well as the following socioeconomic variables: age group (under 70 and 70 and over), gender, psychiatric diagnosis history, education level, employment status, marital status, negative financial impact of the pandemic, risk medical condition. Education, employment status and marital status were all dummy coded. A linear and quadratic term for time since study start and a 2020 indicator variable was added to estimate the effect of the pandemic on PHQ-9 and GAD-7 scores after removing any background trend in previous years. Zero-inflated models did not include an interaction term with 2020 (models were not significantly improved by including an interaction term). Therefore the zero-inflated component does not tell us anything specific about the effect of 2020 so for simplicity they are not reported here. Incidence rate ratios were calculated to illustrate the incremental effect on PHQ-9/GAD-7 scores of each risk factor in 2020 relative those without the risk factor.

All statistically significant variables were included in a final adjusted model to assess independent effects. Predicted values from the adjusted final model were obtained and plotted for year 0 (study start, October 2015), 1 year, 2 years, 3 years after study start, and during the pandemic (i.e. ∼4.5 years after study start).

Of the 3,281 people who completed the COVID-19 mental health questionnaire in 2020, 2,238 had four previous data points; 566 had 3; 415 had 2; and 62 had 1 (Figure 1). The distribution of assessment by month in each year is shown in the supplement.

**Figure 1.**
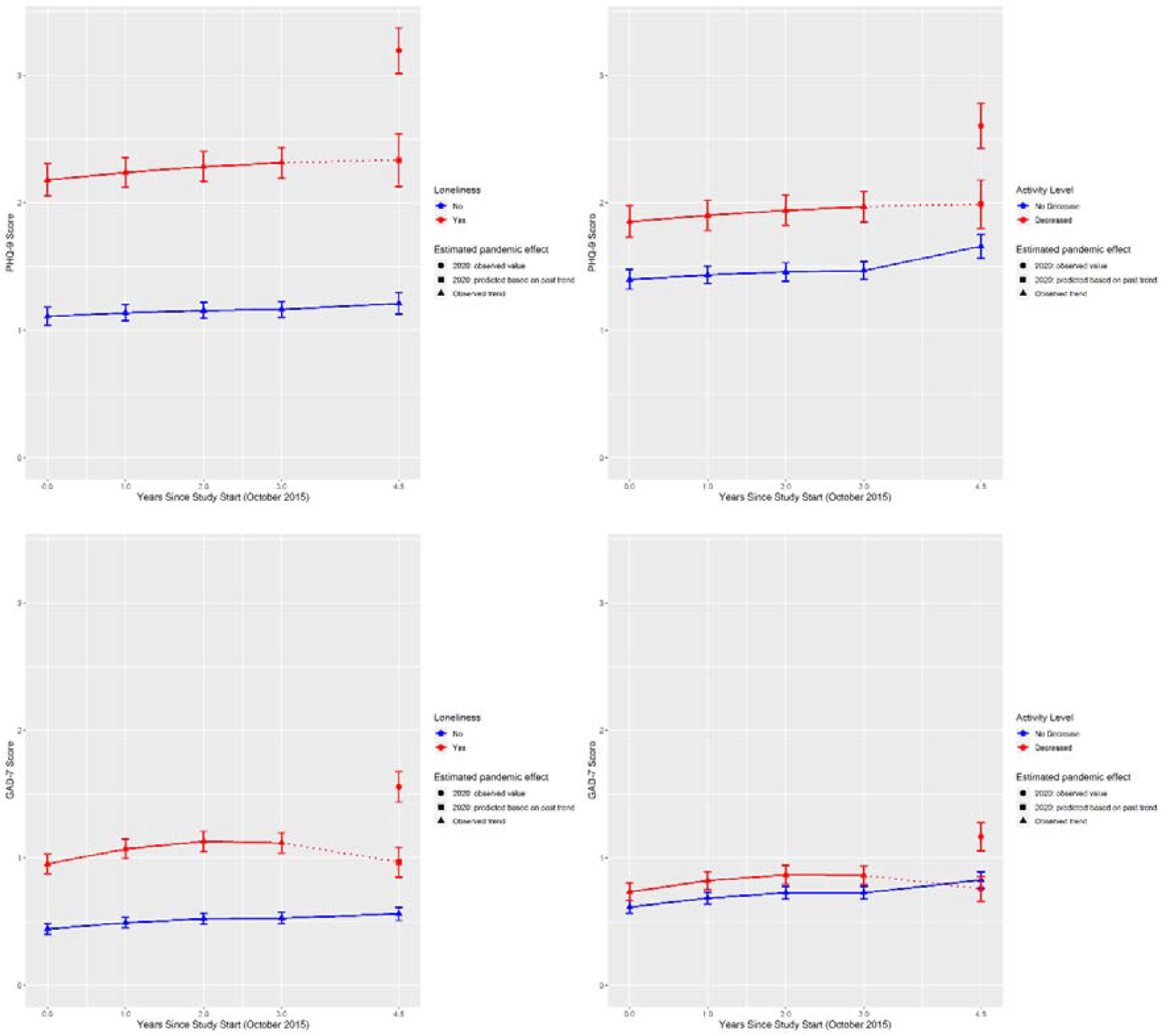
Trajectories of predicted PHQ-9 and GAD-7 scores from zero-inflated negative binomial regression models for loneliness and physical activity.

Statistical analysis was undertaken in the R software environment for statistical computing. Longitudinal zero-inflated negative binomial regression models were fitted using the package glmmTMB (https://github.com/glmmTMB/glmmTMB).

### Role of the funding source

The funder had no role in any part of the project. The corresponding author had full access had full access to all the data in the study and had final responsibility for the decision to submit for publication.

## Results

### Participants

In total, 3,281 people completed the COVID-19 mental health questionnaire, 542 of these either joined PROTECT in May/June 2020 specifically to do the COVID-19 element or they joined PROTECT after the October 2019 data freeze so no pre-pandemic data available for this analysis. These were excluded but there were no major differences in characteristics between the 3,821 used in this analysis and the 542 excluded (see supplement). The characteristics of the sample analysed are described in [Table ***1***. The mean age in 2020 was 67 (standard deviation 6.5, range 55-96), around one third had an undergraduate level education, 80% were female and 98% were white (because of the very low numbers of other ethnicities, ethnicity was not considered further in the analysis). These figures are similar to the wider 25,000 PROTECT study sample (18). Twenty-six (0.7%) people reported having a new continuous cough or high temperature in the last two weeks, a similar proportion (1%) reported a family member with these symptoms. One person in the sample had been hospitalized with COVID-19 in the last four weeks.

**Table 1.** Demographics characteristics for the whole sample.

|  | **Total** | **%** |
| --- | --- | --- |
| **Age Group** |  |  |
| 70 and Over | 1,001 | 30 |
| Under 70 | 2,280 | 70 |
| **Gender** |  |  |
| Female | 2,610 | 80 |
| Male | 671 | 20 |
| **Marital Status** |  |  |
| Married/ Civil Partnership/Co-habiting | 2,421 | 74 |
| Widow/Separated /Divorce | 615 | 19 |
| Single | 245 | 7 |
| **Education** |  |  |
| School to 16 | 400 | 12 |
| 16 to 18 | 1,006 | 31 |
| Undergrad | 1,142 | 35 |
| Post-grad | 733 | 22 |
| **Employment** |  |  |
| Employed (full-time) | 509 | 16 |
| Employed (part-time) | 569 | 17 |
| Self-employed | 280 | 9 |
| Retired | 1,847 | 56 |
| Unemployed | 76 | 2 |
| **Lifetime history of any psychiatric illness** |  |  |
| No | 2,134 | 65 |
| Yes | 1,147 | 35 |

### Risk factors and trajectories

#### Case level analysis

In the cross sectional pandemic data, only variable not associated with higher proportions of mild and moderated-to-severe depressive and anxiety symptoms was education level (see supplement for proportions). Mild and moderate to severe anxiety cases were generally less common and the only variables not associated with higher proportions of cases were education level and marital status.

We then compared case level differences in 2019 with 2020. Overall case level estimates for moderate-to-severe symptoms were comparable across the two years. One hundred and eighty-five (5.6%, 95% CI: 4.9-6.4) and 89 (2.7% 95% CI: 2.2-3.3) of 3,281 people in 2020 had a PHQ-9 score ≥ 10 and a GAD-7 score ≥ 10 respectively. This compared with 124 (4.1%, 95% CI: 3.5-5) and 66 (2.2%, 95% CI: 1.8-2.8) respectively with moderate-to-severe symptoms in 2019 (n=2,959). There was a more pronounced difference in mild symptoms. In 2020 634 (19%, 95% CI: 18-20.7) has mild depressive symptoms compared with 392 (13.2%, 95% CI: 12.1-14.5) in 2018. Similarly, 415 people had mild anxiety symptoms in 2020 (12.6%, 95% CI: 11.6-13.8), compared with 276 in 2019 (9.3%, 95% CI: 8.3-10.4).

#### Trajectories of PHQ-9 and GAD-7 scores

The results from the univariate analysis are shown in supplementary data. Loneliness, decreased physical activity, being a woman and being retired were all associated with significant worsening of depressive symptoms in 2020. Similarly loneliness, decreased physical activity and being a woman were also associated with worsening GAD-7 scores in 2020. Not being in full time employment was associated with a greater worsening of GAD-7 score relative to being full time employed. Both those with a psychiatric history and those without experienced worsening symptoms during the pandemic, but the change was relatively higher in the no history group. The absolute GAD-7 score for people with a psychiatric diagnosis was consistently higher throughout the entire study period.

For the final adjusted model of PHQ-9 trajectory, loneliness, activity level, gender and retirement status were all included as covariates and for the GAD-7 adjusted model, loneliness, physical activity, gender, full time employment status and history of psychiatric condition were included as covariates.

Results from the adjusted models are shown in Table 2 and predicted adjusted PHQ-9 and GAD-7 scores for each time point are provided in full in the supplement, plots of these predicted values for loneliness and physical activity are shown in [Figure ***2***.

**Table 2.** **Adjusted negative binomial regression component of ZINB models of PHQ-9 and GAD-7. Regression coefficients represent the effect of 2020 on scores adjusted for background trends in previous years.**

|  | **PHQ-9** |  |  |  |
| --- | --- | --- | --- | --- |
| **Risk factor** | **IRR** | **L 95% CI** | **U 95% CI** | **P** |
| Loneliness | 1.29 | 1.20 | 1.38 | <0.0001 |
| Activity level decreased | 1.15 | 1.08 | 1.22 | <0.0001 |
| Women | 1.13 | 1.04 | 1.23 | 0.004 |
| Retired | 1.11 | 1.04 | 1.18 | 0.001 |
|  | **GAD-7** |  |  |  |
| Loneliness | 1.25 | 1.13 | 1.38 | <0.0001 |
| Activity level decreased | 1.17 | 1.07 | 1.27 | 0.001 |
| Women | 1.15 | 1.01 | 1.30 | 0.03 |
| Full time employed | 0.91 | 0.81 | 1.03 | 0.15 |
| History of psychiatric condition | 0.84 | 0.77 | 0.92 | 0.0002 |

ZINB: zero-inflated negative binomial regression; IRR: incidence risk ratio

##### Loneliness

Loneliness was associated with a 30% increase in PHQ-9 score in 2020, relative to the not lonely group (IRR=1.3, 95% CI: 1.2-1.4, p<0.0001). Prior to 2020 people rated as lonely scored approximately one point higher than those rated not lonely ([Figure ***2***). In 2020 (4.5 years after study start), however the difference between the two groups was ∼2 points, with PHQ-9 score increasing to 3.2 (95% CI: 3.0-3.4) among those who reported loneliness and remaining stable (1.2, 95% CI: 1.1-1.3) for those not reporting loneliness. In other words, about 50% of the difference in PHQ-9 score between loneliness and no loneliness during the pandemic was accounted for by the general higher burden of symptoms associated with being lonely. For context, this means that in 2020 people who were lonely reported either a new PHQ-9 symptom for several days of the last two weeks or a worsening of an existing symptom to more than half the days in the last two weeks.

For GAD-7, symptoms were 20% worse in those who rated as lonely (IRR=1.2, 95% CI: 1.1-1.4, p<0.0001). Among those with no loneliness, GAD-7 total score was 0.5 across all years (**Error! Reference source not found**.). For those with loneliness, GAD-7 score was 0.5 higher (∼1) in years prior to 2020 but in 2020 the score increased to 1.6 (95% CI: 1.5-1.7). Again, the pandemic accounted for around 50% of the difference in GAD-7 scores attributable to loneliness in 2020.

##### Physical activity

Decreased physical activity during the pandemic was associated with a 20% increase in PHQ-9 and GAD-7 symptoms during 2020 (IRR=1.2, 95% CI: 1.1-1.2, p<0.0001 and IRR=1.2, 95% CI: 1.1-1.3, p<0.0001 respectively). The general trajectory of PHQ-9 and the difference in scores between those reporting a decreased in physical activity, and those not, was similar to loneliness, although the difference was smaller ([Figure ***2***). That is, there was around a 0.5 point difference in the years prior to 2020 and 1 point difference in 2020 (decreased physical activity: 2.6, 95% CI: 2.4-2.8, no decrease: 1.7, 95% CI: 1.6-1.8). Similar to loneliness again, GAD-7 score was modestly higher for people with decreased physical activity in the years prior to 2020 (Figure 2). However in 2020, GAD-7 score increased to 1.2 (95% CI 1.1-1.3) among those reporting decreased physical activity, which compares with 0.9 for those with no decrease in physical activity (95% CI: 0.8-0.9).

##### Gender, employment status and psychiatric history

Being a woman and being retired were both also associated with worsening PHQ-9 symptoms during 2020 (IRR=1.1, 95% CI: 1.0-1.2, p=0.004; IRR=1.1, 95% CI: 1.0-1.2, p=0.001). Similarly, being a woman was also associated with worse GAD-7 scores in 2020 (IRR=1.2, 95% CI: 1.0-1.3, p=0.03). Having a history of a psychiatric condition was associated with a relatively lower score in 2020 (IRR=0.8, 95% CI: 0.8 -0.9, p=0.0002). However, this was driven by an increase in mental health symptoms in those with a history of psychiatric diagnosis. The absolute levels GAD-7 score for people with a psychiatric history remained higher in 2020 than those without (1.44 [95% CI: 1.3-1.59] vs 0.69 [95% CI: 0.63-0.75], see supplement). The association between full time employment and improved mental health identified in the univariate analysis did not hold in the adjusted analysis (Table 2).

## Discussion

This is the first longitudinal study to focus specifically on the links between loneliness, physical activity and mental health during the COVID-19 pandemic with longitudinal data pertaining to pre-pandemic mental health. Overall, in a cohort aged between 55 and 96, there was an increase in the proportion of people with mild depressive symptoms from 13.2% in 2019 to 19% in 2020 and an increase in the proportion of people with mild anxiety symptoms (from 9.3% to 12.6%). Proportions of people with moderate-to-severe symptoms were comparable. Longitudinally, both loneliness and decreased physical activity were associated with worse mental health in 2020 compared to previous years, suggesting that the association observed in 2020 was not due to a longer standing relationship between current loneliness and physical activity and mental health before 2020. Our data also show that the impact of the pandemic on mental health would be overestimated without the longitudinal perspective, bringing new insight to these established mental health risk factors and in line with recent findings related to other risk factors (22).

Around half of the sample reported some degree of loneliness during the pandemic. Loneliness during the pandemic was associated with a 1 point higher score on the PHQ-9 between 2015 and 2019 compared to people who did not report loneliness, but this difference doubled to 2 points during the pandemic. In contrast, there was no worsening of mental health symptoms for people who did not report loneliness. Over one third of the sample reported decreased physical activity during the pandemic. The effect on PHQ-9 scores was more modest that of loneliness but was nevertheless associated with a worsening of symptoms during the pandemic. There were also statistically significant increases in GAD-7 scores for both loneliness and decreased physical activity though the differences were smaller than for PHQ-9 scores. Collectively, these findings emphasise the potential impact of finding novel solutions to tackle loneliness and decreased physical activity during the pandemic and underscore the important general relationship between the two and mental health (13,16,23).

Of the socioeconomic and demographic variables analysed, both being a woman and being retired were associated with pandemic-specific worsening in mental health, in line with previous UK representatively sampled studies (9,24). While our data do not show any increase in mental health symptoms related to other socioeconomic factors specifically associated with the pandemic (e.g. rating that the pandemic had a negative financial impact or employment status) we believe it would be premature to rule out an effect, firstly because the economic impact of the pandemic has not yet fully taken hold and secondly because we note other large representative surveys have reported clear links (8,9). Finally, similar to other studies we found no evidence that having a medical condition associated with increased risk of severe COVID-19 was associated with worsening symptoms of depression and anxiety (9).

### Limitations

One important limitation is the potential for bias in an on-line self-selecting sample. In particular we note the overrepresentation of women, white British people and those with a higher education, however because our analysis is focused on longitudinal patterns rather than prevalence there is still merit in identifying these trends within this sample. A second limitation is determining causation, a pervasive issue observational studies. Because the loneliness and physical activity questions were only taken during the pandemic it may be the case that worse mental health drove a decrease in physical activity and increase in loneliness. The wider literature has highlighted a causal relationship between higher physical activity levels and lower risk for major depressive disorder (but no causal relationship for the reverse) so in the context of this evidence it would be reasonable to hypothesise that maintaining physical activity during the pandemic may mitigate risk of mental health deterioration (25). A large randomised control trial would be needed to assess this but our findings pave the way for robust intervention testing. We are not aware of any studies which have conclusively shown a causal directional link between loneliness and mental health but the well-established link between the two is one of the reasons why loneliness is a critical policy area in the UK and internationally. Here we are able to show that for the first time that the association between loneliness and worse mental health is not solely due to those who are currently lonely having long-standing worse mental health; there is a specific effect of 2020 in this sample which is an important advance over previous cross sectional studies.

In conclusion, in this large longitudinally studied sample exploring mental health effects of the COVID-19 pandemic in middle aged and older people in the UK, we found that loneliness and decreased physical activity were both associated with worse mental health during the pandemic and that this was distinct from the general relationship between these two risk factors and poor mental health. Our study provides robust evidence in support of targeted interventions to improve mental health of people in mid to late life in the subsequent waves of the pandemic.

## Data Availability

The data that support the findings of this study are available but restrictions apply to the availability of these data, which were used under license for the current study, and so are not publicly available. Data are however available from the authors upon reasonable request and with permission of the PROTECT study.

## Declaration of interests

The other authors declare no competing interests. The views expressed are those of the author(s) and not necessarily those of the NHS, the NIHR or the Department of Health and Social Care or Public Health England.

## Funding

This paper represents independent research part funded by the National Institute for Health Research (NIHR) Biomedical Research Centre at South London and Maudsley NHS Foundation Trust and King’s College London. This research was also supported by the NIHR Collaboration for Leadership in Applied Health Research and Care South West Peninsula. Additional funding for the COVID-19 mental health questionnaire was provided by King’s College London and Stavanger University Hospital.

## Acknowledgements

The authors would like to thank Ellie Pickering, Adam Bloomfield and Ben Wood at the University of Exeter for project management and IT development related to the COVID-19 mental health questionnaire. We also thank the HRA for expediting ethical review, the PROTECT study PPI group and all participants for taking the time to complete the questionnaire. Dr Siobhan O’Dwyer is supported by the National Institute for Health Research Applied Research Collaboration South West Peninsula. The views expressed in this publication are those of the author(s) and not necessarily those of the National Institute for Health Research or the Department of Health and Social Care.

## Author contributions

BC: design, analysis, figures, literature search, data collection, interpretation, drafting, review; ZK: design, funding, analysis, manuscript drafting and review; WH: statistical analysis and data visualisation, manuscript review; SOD: design, manuscript drafting and review; AC: data collection; MVS: funding, data collection; KM: literature search; NW: design, manuscript drafting and review; IT: design, funding, manuscript drafting and review; DA: design, funding, manuscript drafting and review; CB: design, funding, manuscript drafting and review.

## Data availability

The data that support the findings of this study are available on request from but restrictions apply to the availability of these data, which were used under license for the current study, and so are not publicly available. Data are however available from the authors upon reasonable request and with permission of the PROTECT study.

---

**Figure 1.**
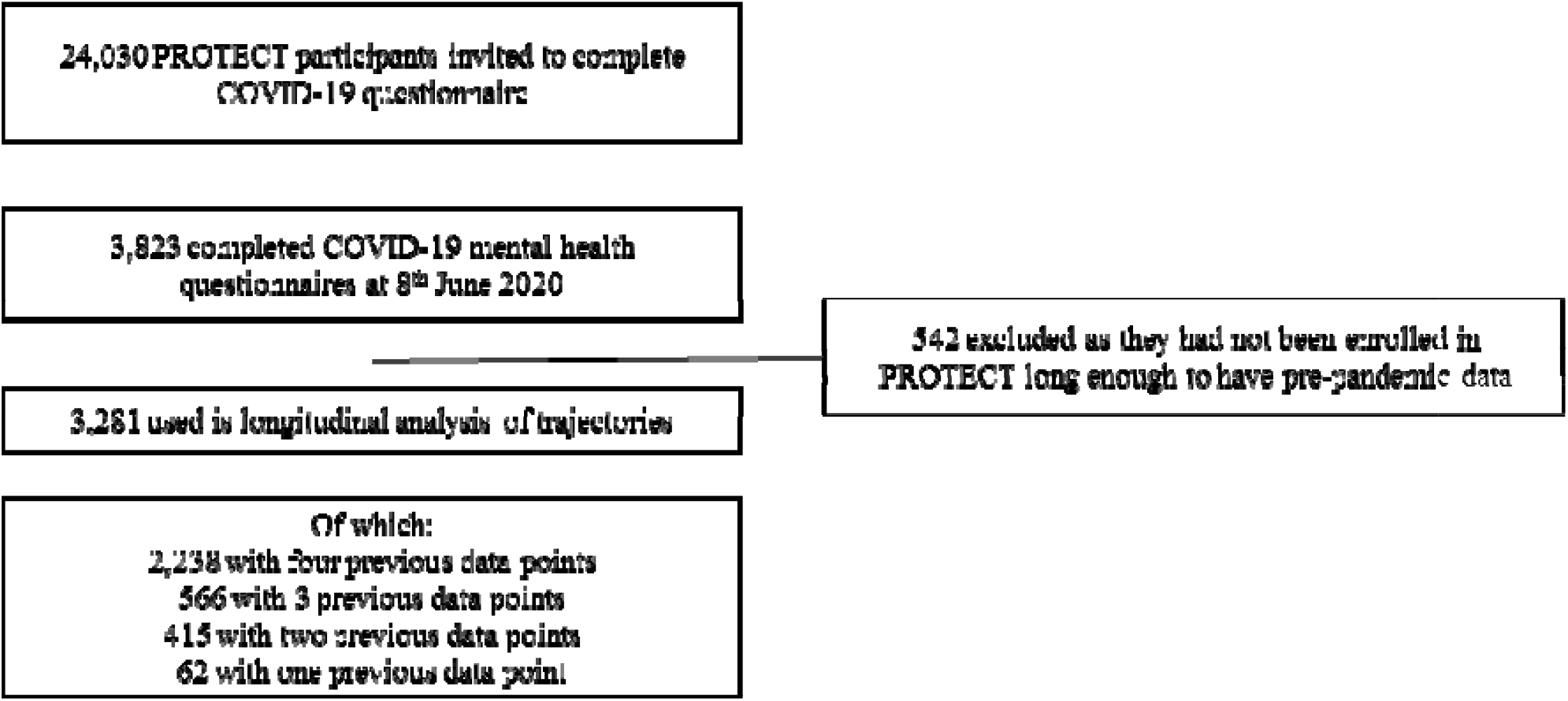
Selection flow chart.

## Notes

### Competing Interest Statement

The authors have declared no competing interest.

### Author Declarations

The PROTECT study received ethical approval from the UK London Bridge National Research Ethics Committee (Ref: 13/LO/1578) and the COVID-19 mental health questionnaire was approved by the same committee (as an amendment) on 6th April 2020.

### Summary of Updates

Add author Siobhan O'Dwyer who was omitted in error during submission (name present on the pdf file)

